# Association between time spent with family and loneliness among Japanese workers during the COVID-19 pandemic: a cross-sectional study

**DOI:** 10.1101/2021.09.30.21264346

**Authors:** Rintaro Fujii, Yusuke Konno, Seiichiro Tateishi, Ayako Hino, Mayumi Tsuji, Kazunori Ikegami, Masako Nagata, Reiji Yoshimura, Shinya Matsuda, Yoshihisa Fujino, for the CORoNaWork Project

## Abstract

**Background:** The current coronavirus (COVID-19) pandemic has had large impacts on society, including people practicing social distancing. This behavioral response has increased loneliness. Loneliness not only increases the risk of psychiatric disorders, but also affects occupational mental health. To avoid the negative effects of isolation, it is important to have social contact with other people, especially family members. Employment and economic instability caused by COVID-19 may have also affected family relationships. It is important to understand the association between family relationships and loneliness in workers under the pandemic.

**Methods:** We collected usable data from 27,036 Japanese workers who completed an online survey during the COVID-19 pandemic. Participants were asked how long they spend with members of their family during mealtimes or at home, and if they experienced loneliness; the latter was assessed by a single question. Other questions included whether participants lived with their spouse, or with someone in need of care. To estimate the odds ratios (ORs) of time with family associated with loneliness we used a multilevel logistic model nested in the prefecture of residence, with adjustments for age, sex, marital status, presence of a cohabitant requiring care, equivalent income, educational level, frequency of remote work, availability of someone for casual chat, smoking, drinking, time for leisure interests, and cumulative rates of COVID-19 in the prefecture.

**Results:** Ten percent (2,750) of the 27,036 participants reported loneliness. The survey showed a significant negative correlation between time spent with family and loneliness (*p*<0.001): participants who spent more time with family were less likely to feel loneliness. In addition, not living with a spouse and living with someone in need of care were associated with loneliness (not living with a spouse: *p*<0.001; living with someone in need of care: *p*<0.001).

**Conclusion:** Loneliness under COVID-19 pandemic conditions was negatively associated with time spent with family members, with the converse result found for participants cohabiting with someone in need of care. These associations suggest the potential value of changes to working practices and interventions to combat loneliness.

## 1 Introduction

The coronavirus (COVID-19) outbreak caused by SARS-CoV-2 in December 2019 has resulted in a global pandemic that has led to multiple public health issues related to mental health (1). COVID-19 is highly infectious and can lead to serious illness, so efforts to prevent the spread of the disease have been implemented worldwide. The strongest effort was to lock down cities and restrict human movements and contacts. In addition, WHO recommended avoiding the “Three Cs,” namely closed spaces, crowded places and close-contact settings, to minimize transmission of the disease. Furthermore, to restrict people’s movements, the Japanese government requested companies to implement remote work, which many companies urgently adopted. Remote work reduces opportunities to communicate with workmates and to receive support from the workplace (2). Although these infection control measures are considered effective in preventing the spread of disease, they also potentially increase the risk of loneliness and mental health problems, both of which have become new public health challenges (3).

Loneliness is defined as “a distressing feeling that accompanies the perception that one’s social needs are not being met by the quantity or especially the quality of one’s social relationships’’ (4). Biologically, it has been linked to activity in the ventral striatum and parietal junction (5), while epidemiologically it has been linked to social status factors such as education, and income (6,7). Loneliness is associated not only with psychological distress (8,9), but also depression and anxiety (10,11), sleep disorders (12), alcoholism (13), Alzheimer’s disease (14), and other psychiatric disorders. Loneliness is also associated with increased mortality and suicidal ideation (15,16). In Japan, suicides attributed to loneliness have increased rapidly since the pre-COVID-19 pandemic period (17). Loneliness may influence not only mental health in general, but also occupational mental health. For example, workers with loneliness are more likely to feel low job satisfaction and express frustration (18).

To avoid isolation, it is important to have contact with other people. It is generally considered that people have the highest frequency of contacts with family members (19). Family involvement is not only related to frequency of social contacts, but also to reported levels of happiness (20). People living alone reported more loneliness than those living with others (21). Another study showed that people without partners, such as divorcees and widows, were more likely to have loneliness (11). On the other hand, people living with family members in need of care are more susceptible to feeling stressed due to the burden of care (22). Therefore, it can be hypothesized that spending time with close family members may reduce loneliness, but this effect may vary depending on the family situation.

We hypothesized that workstyle changes resulting from the COVID-19 pandemic, such as remote work, have resulted in reduced opportunities to interact with others and have led to increased loneliness among workers. At the same time, as remote work continues to increase due to the COVID-19 pandemic, workers are released from commuting time and therefore have more time to spend with their families. In fact, before COVID-19, remote work was recommended in Japan from the perspective of work-life balance. However, employment and economic problems caused by COVID-19 may have also affected family relationships. Therefore, this study aimed for a better understanding of the association between family relationships and loneliness in workers during the COVID-19 pandemic.

## 2 Methods

### 2.1 Study Design and Participants

We conducted an online survey from December 22 to 26, 2020, under the Collaborative Online Research on the Novel-coronavirus and Work (CORoNaWork) Project. Information about the protocol for this cross-sectional study has already been published (23). The target population comprised workers with a full-time employment contract at that time. In total, 33,087 participants completed the survey, and once invalid answers were excluded, data from 27,036 were available for analysis. The study was approved by the ethics committee of the University of Occupational and Environmental Health, Japan. Participants provided informed consent by filling out a form on the survey website.

### 2.2 Assessment of time spent with family during mealtimes or at home

The survey included questions designed to find out how much time participants spend with their family for meals or simply at home. To the question: “How long do you spend with family having a meal or at homeã” participants selected one of the following options: more than 2 hours, more than 1 hour, more than 30 minutes, less than 30 minutes, and almost never. The following questions were also included in the survey, and required “Yes” or “No” answers: “Do you live with your spouseã” and “Do you live with someone in need of careã”

### 2.3 Assessment of loneliness

One question focused on whether the participants experienced loneliness or not. To the question: “During the last 30 days, how frequently have you felt lonelinessã” participants selected one of the following options: never, a little, sometimes, usually, always. Answers “always,” “usually,” or “sometimes” were taken as indicating loneliness.

### 2.4 Other covariates

The following demographic and socioeconomic factors were included as covariates: age, sex, marital status, presence of a cohabitant in need of care, equivalent income, educational level, frequency of remote work, presence of someone for casual chat, smoking, drinking, time for leisure interests and cumulative rates of COVID-19 in the prefecture of residence.

The cumulative incidence of COVID-19 in the prefecture of residence between the time of the survey and one month later was used as a community-level variable. The relevant information was obtained from public institution websites.

### 2.5 Statistical analysis

We used a multilevel logistic model to estimate odds ratios (ORs) for time spent with family during mealtimes or at home and loneliness. Loneliness was identified only if participants answered always, usually, or sometimes to that question. The multivariate model was adjusted for the factors: age, sex, marital status, presence of a cohabitant requiring care, equivalent income, educational level, frequency of remote work, presence of someone for casual chat, smoking, drinking, time for leisure interests and cumulative incidence rate of COVID-19 in the prefecture.

P-values below 0.05 were considered statistically significant. All analyses were run on Stata Statistical Software Release 17. (StataCorp LLC, College Station, TX, USA.).

## 3 Results

Table 1 shows the basic characteristics of the participants. Ten percent (2,750) of the 27,036 participants experienced loneliness. When asked about time spent with family, the largest group (26.6%) answered “almost never;” however, among participants with loneliness, 46.1% answered “almost never” to this question. Of those who spent more than 2 hours with family, 9.9% had loneliness. Notably, the incidence of loneliness decreased as the time spent with family increased. In addition, loneliness was reported less frequently by participants who lived with their spouse, but more frequently by those living with someone in need of care.

**Table 1.**
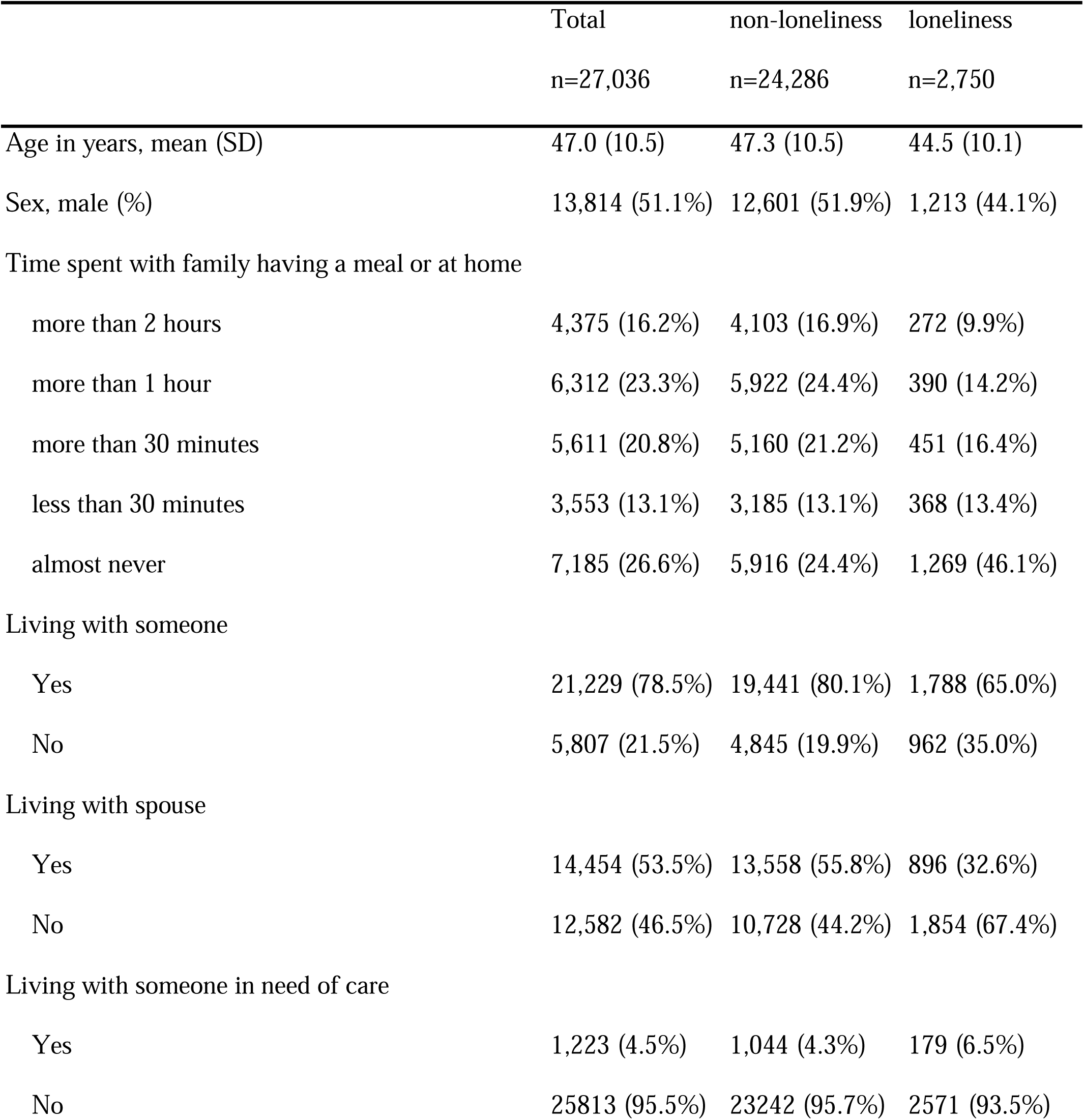
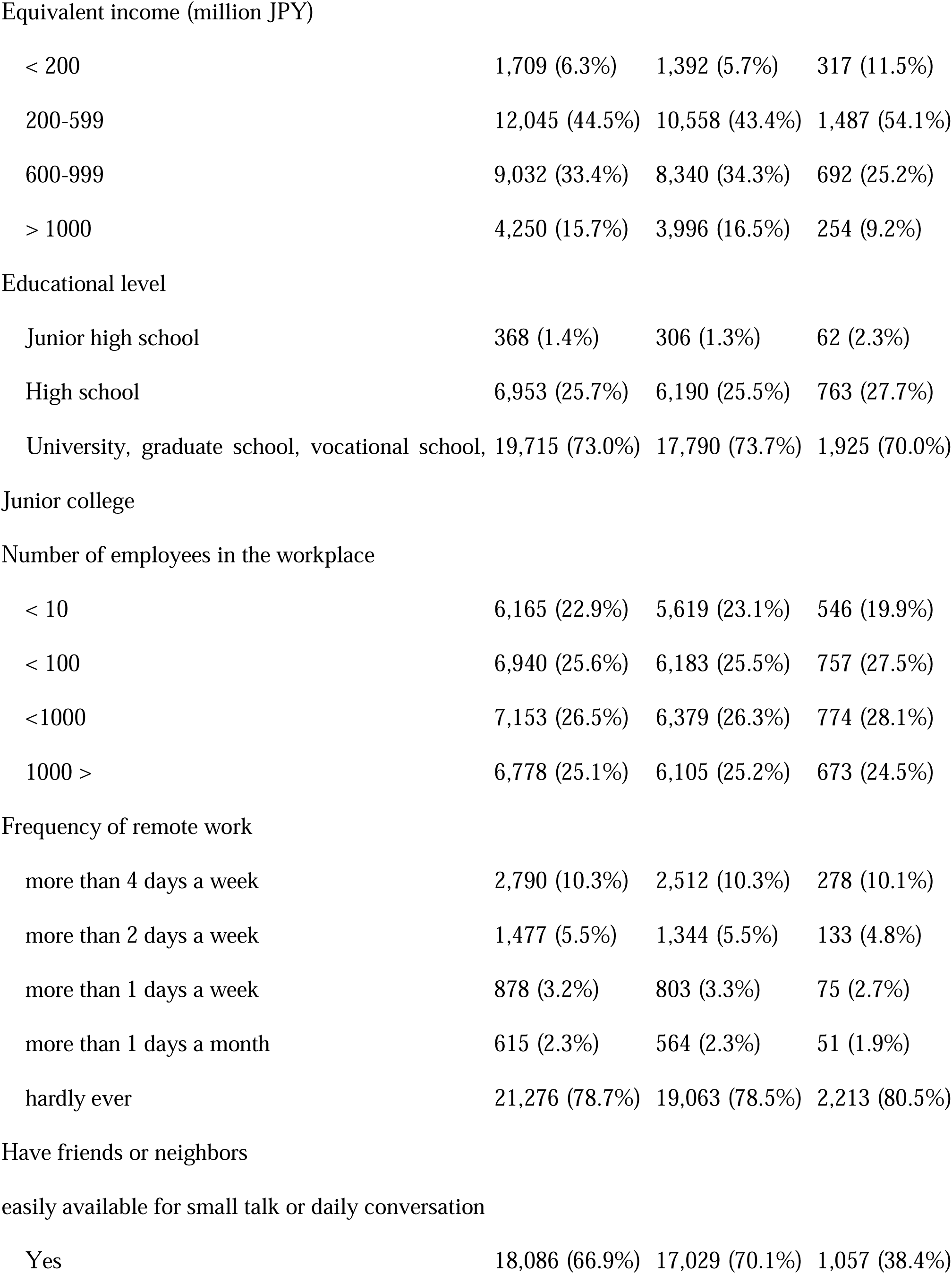
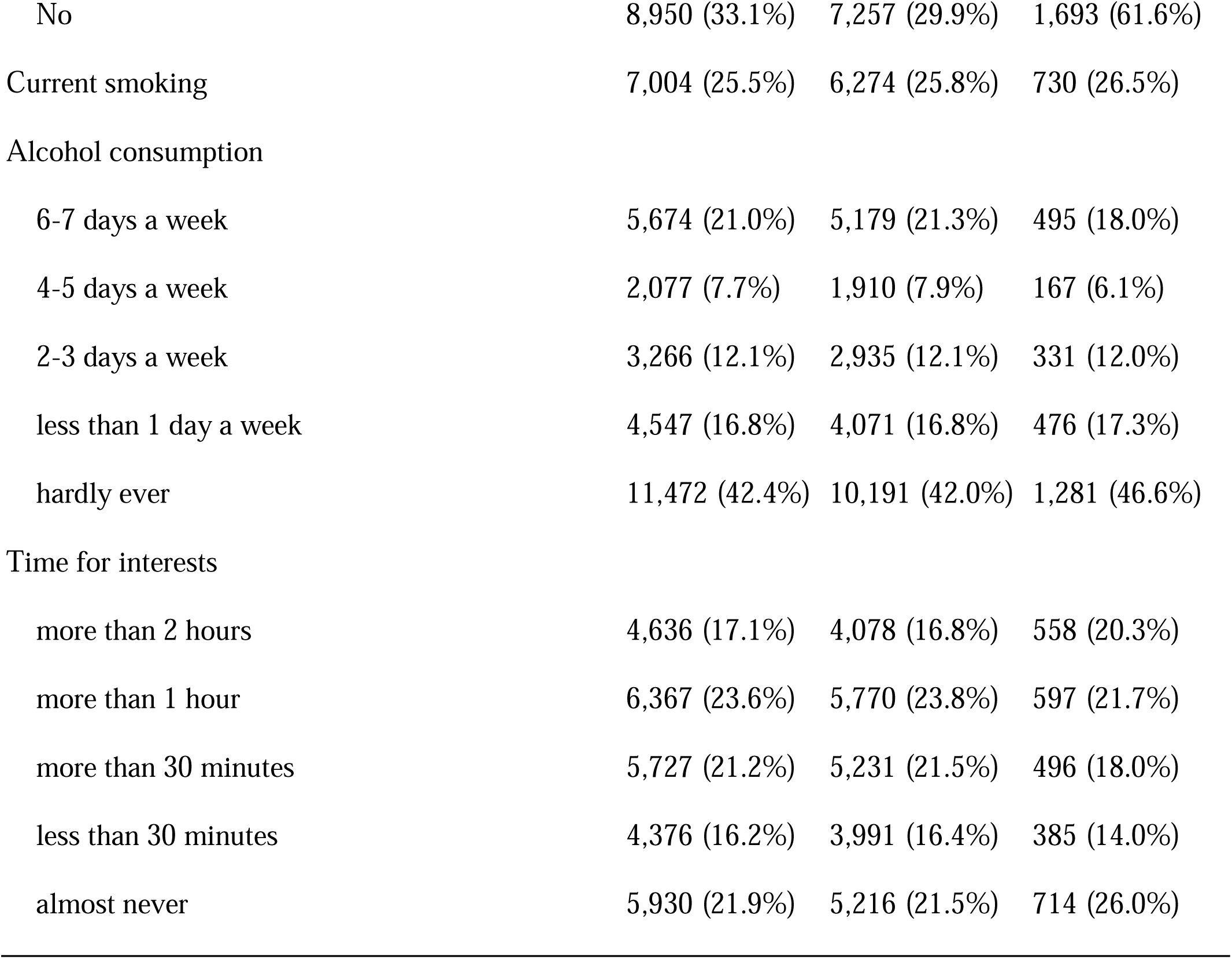
Characteristics of participants who experienced loneliness.

Table 2 shows the odds ratio (OR) of time spent with family and loneliness estimated by the logistic model. The age-sex adjusted OR of loneliness for those who reported spending little or no time with family indicated a significant association (OR=3.43, 95% CI 2.99-3.94, *p*<0.001). This result was similar with multivariate analysis (OR=2.00, 95% CI 1.71-2.33, *p*<0.001). In addition, not living with a spouse and living with someone in need of care were associated with loneliness (OR=2.44, 95% CI 2.24-2.67, *p*<0.001, and OR=1.67, 95% CI 1.41-1.97, *p*<0.001, respectively). Again, similar results were obtained with multivariate analysis (not living with a spouse: OR=1.44, 95% CI 1.30-1.61, *p*<0.001; living with someone in need of care: OR=1.85, 95% CI 1.56-2.21, *p*<0.001).

**Table 2.**
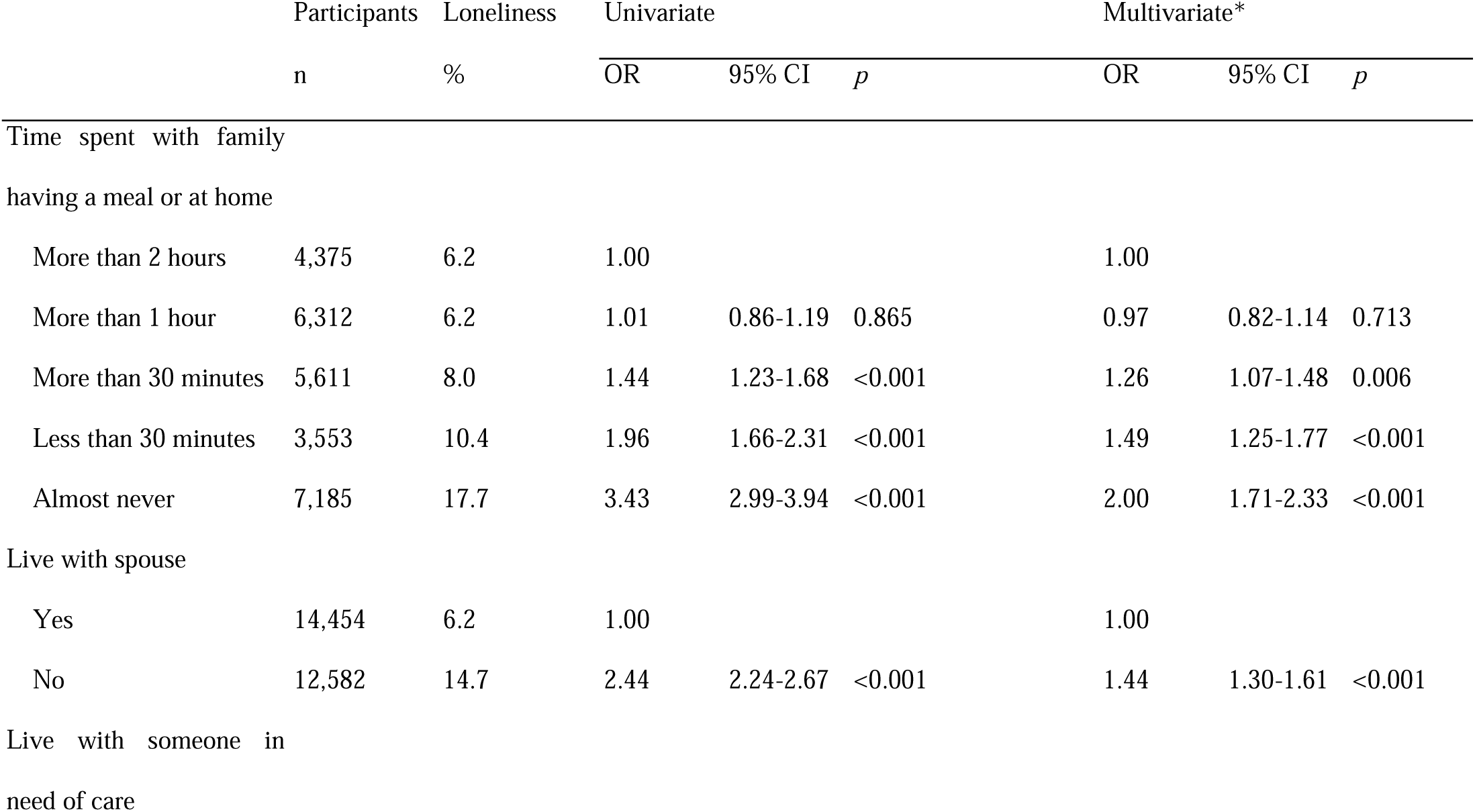
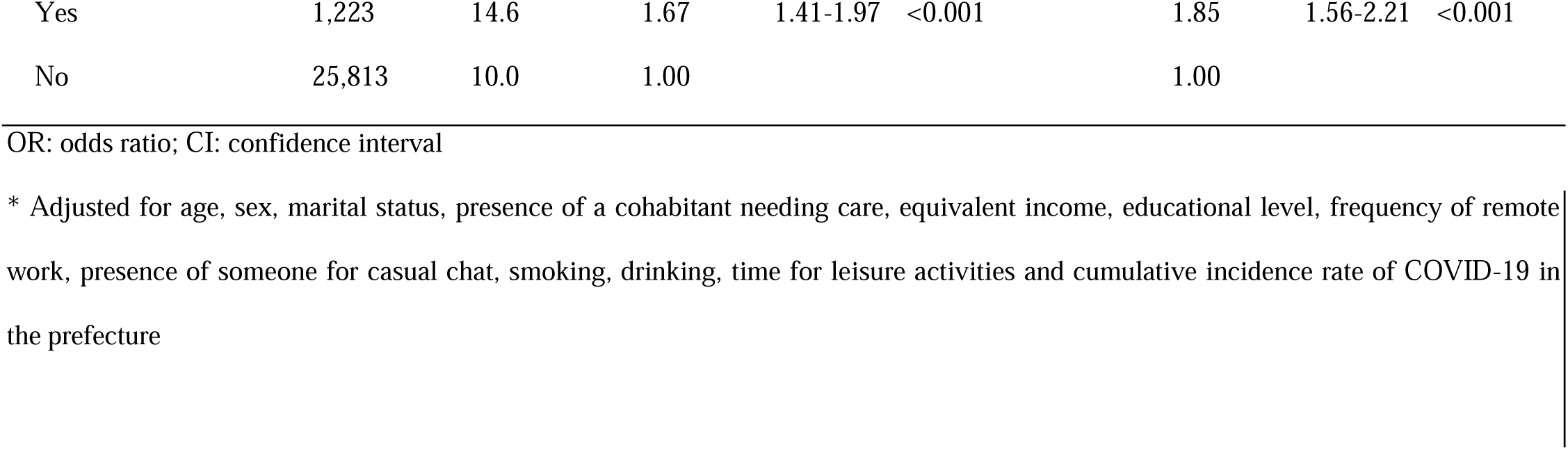
Association between time spent with family having a meal or at home, and loneliness.

## 4 Discussion

This study showed that during the COVID-19 pandemic, workers in Japan who spent less time with their families were more likely to report loneliness. Those who did not live with their spouse were also more likely to feel lonely than those who did live with their spouse. However, time spent with family members in need of care and attention was associated with more loneliness.

The relationship between time spent with family and loneliness showed a dose-response function: the shorter the time spent with family, the greater the likelihood of feeling lonely. These results were robust even after adjusting for factors such as income, social status, and education, which have been reported to be associated with loneliness in previous studies. Time spent with family members is thought to contribute to well-being because of the social integration provided by the family, enhanced importance of the self, and the accessibility of social support (24). Low well-being is correlated with loneliness (25), possibly further illustrating the relationship between time spent with family and loneliness. Previous studies have also reported that living without a partner is a contributory factor in loneliness (26), while others have suggested that not only cohabitation but also the strength of the relationship between partners modulates the intensity of loneliness (27). Our results provide evidence in support of the importance of strong family relationships, as the less time spent with family, the more participants were likely to report loneliness.

Biological mechanisms have been proposed to explain the association between time spent with family and loneliness. Dopamine can be involved in loneliness. Dopaminergic nerves serve in regulating reward-processing behavior mediated by pleasure and enjoyment, and emotional behaviors such as romantic love. The ventral striatum is a neurotransmitter-related region centered on dopamine and may be involved in feelings of loneliness (5). This dopaminergic function may mediate loneliness (28). In a survey of female workers on how they spend their time in daily activities, most participants described time spent with their spouse and family as enjoyable, whereas time alone was not enjoyable (29). A similar association between loneliness and family time was found in this study. Because this study assessed loneliness using the subjective question “How frequently have you felt lonelinessã”, the results seem likely to be influenced by subjective experiences such as pleasure and enjoyment. We presume that dopamine nerves of the reward system are implicated in the finding that the less time spent with family, the more loneliness participants experienced.

Interestingly, loneliness varied with factors other than simple time spent with family. In contrast to living with a spouse, living with a person in need of care was strongly positively correlated with loneliness. The burden of care on family members increases not only psychological stress but also economic burden (30). A previous study reported that caring for a family member was correlated with loneliness particularly when the family members lived together (31). Research has also suggested that emotional support from others, social connections and contact with friends are important for countering caregivers’ feelings of loneliness (32,33).

This study prompts two suggestions. First, time spent with family may be useful in reducing loneliness, as the mental health impact of the COVID-19 pandemic becomes a global public health issue. Second, recommended measures for preventing COVID-19 infection, such as maintaining social distance and refraining from going out, may lead to reduced opportunities for communication with others, which may in turn negatively affect mental health (3). However, increased time spent with family due to increased time spent at home, such as in the case of remote work, may compensate for the effects of reduced opportunities for socializing. Furthermore, home-alone and single workers are a high-risk group for loneliness, and may require careful support in terms of mental well-being.

Several limitations of the present study should be addressed. First, the results are based on an Internet survey, and so the generalizability of the results is open to question. However, we purposively aimed for a diverse target population in terms of gender, occupation, and region, based on COVID-19 incidence data. Second, although there are several ways to assess loneliness (34), we did so by using a single question, following previous studies on loneliness that used a single assessment item (35). Finally, the survey was conducted during the COVID-19 pandemic, but it remains unclear how the pandemic and resulting changes in daily and occupational environments might have affected the survey outcomes.

In this study, 10% of participants reported feelings of loneliness during the COVID-19 pandemic. The feeling of loneliness was associated with time spent with family members, with contrasting results depending on the status of the cohabitants. Greater consideration should be given to interventions such as support for caregivers, and encouragement regarding remote work and other potential changes, consistent with the worker’s family and living conditions.

## Data Availability

Data not available due to ethical restrictions

## Author contributions

Y.F. was the chairperson of the study group. R.F. conceived the research questions. All authors designed the research protocol and developed the questionnaire. R.F. conducted the statistical analysis with Y.F. R.F. drafted the initial manuscript. All authors revised and approved the final manuscript.

## Ethics Statement

The study, involving human participants, was reviewed and approved by the Ethics Committee of the University of Occupational and Environmental Health, Japan (reference No. R2-079 and R3-006). Participants provided informed consent by filling out a form on the survey website.

## Data Availability Statement

Data not available due to ethical restrictions.

## Acknowledgements

We thank other members of the CORoNaWork Project: Akira Ogami, Arisa Harada, Hajime Ando, Hisashi Eguchi, Kei Tokutsu, Kosuke Mafune, Kyoko Kitagawa, Ning Liu, Rie Tanaka, Ryutaro Matsugaki, Tomohiro Ishimaru, and Tomohisa Nagata.

